# High Response Rate to Antidepressants in the Management of Paroxysmal Hypertension (Pseudopheochromocytoma)

**DOI:** 10.1101/2021.09.10.21261667

**Authors:** Samuel J. Mann, Kaushal V. Solanki

## Abstract

There is no widely recognized preventive treatment for patients with paroxysmal hypertension (“pseudopheochromocytoma”) who suffer recurrent and often severe paroxysmal elevation of blood pressure, repeated emergency room visits and hospitalizations. In this case series we assessed the effectiveness of treatment with an antidepressant in preventing recurrent paroxysms. A secondary exploratory objective was to examine the psychological profile of patients and its relationship to response to treatment. The charts of 52 patients who reported having experienced at least 3 symptomatic episodes were selected; and and for whom treatment data were included. Treatment with an antidepressant was offered to, and follow-up data were available in 37..

Two patients refused treatment, 6 were unable to tolerate an effective dose, 3 were lost to follow-up, and 1 was non-compliant with the medication. Of the remaining 25 evaluable patients, 92% (23/25) responded, with 80% (21/25) achieving complete and persisting cessation of paroxysms, and 8% (2/25), a reduction in frequency. Importantly, an antidepressant was effective in nearly all patients who reported that they were not suffering from anxiety or depression. The data were insufficient to determine superiority of any one antidepressant versus another. We conclude that treatment with an antidepressant is effective in preventing hypertensive paroxysms in a high proportion of patients with paroxysmal hypertension. Given the absence of any other pharmacologic intervention capable of preventing recurrent paroxysms, the similarly high response rate observed in previous reports, and the relatively safe profile of antidepressant agents, the findings support more widespread use of an antidepressant in patients with this disorder to prevent years of continued hypertensive paroxysms and their consequences.

## Introduction

Patients with the dramatic syndrome of paroxysmal hypertension (pseudopheochromocytoma) experience episodes of hypertension characterized by sudden and marked blood pressure elevation accompanied by symptoms such as headache, flushing, and diaphoresis. Paroxysms are typically unprovoked, are not related to current stress or emotional distress, and described by patients as occurring “out of the blue.”^1^ Paroxysmal hypertension is observed in approximately 45% of patients with a catecholamine-producing pheochromocytoma, and always generates suspicion of that tumor.^1,2^ However, a pheochromocytoma is found in only 1-2% of patients with paroxysmal hypertension.^1^ In the remaining 98-99%, the cause and treatment have long been an enigma, with most patients remaining ineffectively treated and at risk of repeated emergency room visits, hospitalizations and chronic disability.

In both a reported case series, and a prospective trial, treatment with an antidepressant was reported to be effective in preventing recurrent paroxysms.^3^ .^4^ This report presents a case series that further examines the effectiveness of antidepressants in preventing recurrent paroxysms, and, given their effectiveness, explores underlying psychosocial factors.

## Methods

A search of medical records approved by the Weill Cornell Medicine Institutional Review Board was conducted to examine the records of all patients whose initial visit occurred after the EPIC electronic medical record system was initiated in 2004. All charts with a recorded diagnostic term of “paroxysmal hypertension” were identified (there is no ICD10 code for paroxysmal hypertension). In all cases the presence of a pheochromocytoma was ruled out by appropriate biochemical testing.

In identifying patients with paroxysmal hypertension, it is necessary to differentiate this disorder from the more commonly occurring entity of “labile hypertension.” The latter differs in that blood pressure elevation usually occurs at times of stress, with patients acknowledging emotional distress; usually it is asymptomatic or only mildly symptomatic.^5^ However, no formal clinical criteria have been proposed to define labile hypertension.

To define and further differentiate paroxysmal hypertension (pseudopheochromocytoma) from labile hypertension, the following clinical criteria, based on the previously reported clinical features, were proposed, and were utilized in identifying patients with paroxysmal hypertension included in this case series:

1. Recurrent episodes characterized by the *sudden* (not gradual) onset of blood pressure elevation.
2. Episodes were symptomatic, with a similarly sudden onset of at least one symptom such as headache, palpitations, diaphoresis, flushing, and/or others.
3. The patient insisted that the episodes occurred without provocation, and were not precipitated by stress or anxiety.

The blood pressure level reached during paroxysms was not a criterion, given the absence in many patients of standardized recorded measurements during episodes as well as the marked blood pressure elevation that is frequently incidentally observed in emergency rooms. However, in nearly all cases systolic blood pressure above 180 mmHg was reported during home measurements.

The charts of 221 patients were identified and reviewed; 189 met the three proposed criteria. In order to eliminate patients in whom hypertensive paroxysms might have been a transient phenomenon that resolved spontaneously, the charts of 123 patients who had experienced fewer than three episodes were excluded, leaving 66 charts. An additional 14 patients, who were seen only once or twice without any further follow-up were also excluded, leaving 52 patients.

Charts were searched for the following characteristics recorded at the initial visit: demographic data, physical symptoms accompanying paroxysms, duration and frequency of paroxysms, and medications.

Although there was no specific treatment protocol, all patients were initially given prescriptions for clonidine and/or a benzodiazepine (usually alprazolam) for use as needed at the time of paroxysms. Patients who found this intervention to be inadequate due to persisting frequency of paroxysms, severity of blood pressure elevation or symptoms, or disruption to routine functioning caused by recurring paroxysms, were offered treatment with an antidepressant. Patients who reported no further paroxysms during treatment with an antidepressant were considered to have had a complete response. Patients who reported a substantial reduction in frequency of paroxysms were considered to have had improvement.

In addition to the results of treatment, for exploratory purposes, data from the recorded psychosocial history were reviewed. The presence or absence of self-reported anxiety or depression was noted. Information concerning a past history of abuse or trauma, or untimely loss of a parent or spouse was noted.

## Results

Of the 66 patients, 49 (74%) were female. Mean age at the time of onset was 55 years (age range 40-70).

Table 1 summarizes the description of the paroxysms. In most patients, the duration of paroxysms ranged from 30 minutes to 3 hours. Paroxysms lasted as long as a day in 8 patients. In addition to the physical symptoms, many reported feeling profound fatigue when paroxysms ended, often lasting the rest of the day. Most reported Emergency Room visits, and 30% reported having been hospitalized at least once because of a hypertensive episode.

**Table 1:** Clinical characteristics.

| **Clinical Characteristics** | **Number of patients** |
| --- | --- |
| ➤ Usual Duration of episodes |  |
| • < 30 mins | 10 |
| • 1-3 hrs | 25 |
| • 3-6 hrs | 6 |
| • 6 hrs-1 day | 8 |
| • Variable | 3 |
| ➤ Symptoms during episode |  |
| • Headache | 15 |
| • Chest pain | 8 |
| • Tachycardia | 4 |
| • Diaphoresis | 5 |
| • Flushing | 12 |
| • Shortness of Breath | 5 |
| • Weakness | 2 |
| • Palpitations | 2 |
| • Lightheadedness | 6 |
| ➤ Fatigue or washed out after episodes | 35 |

### Response to treatment

Table 2 summarizes the response to treatment of the 52 patients for whom follow-up data were available. All patients were initially prescribed clonidine and/or alprazolam for the acute management of paroxysms. Thirteen (25%) were satisfied with the effect of this intervention and did not seek other interventions.

**Table 2:**
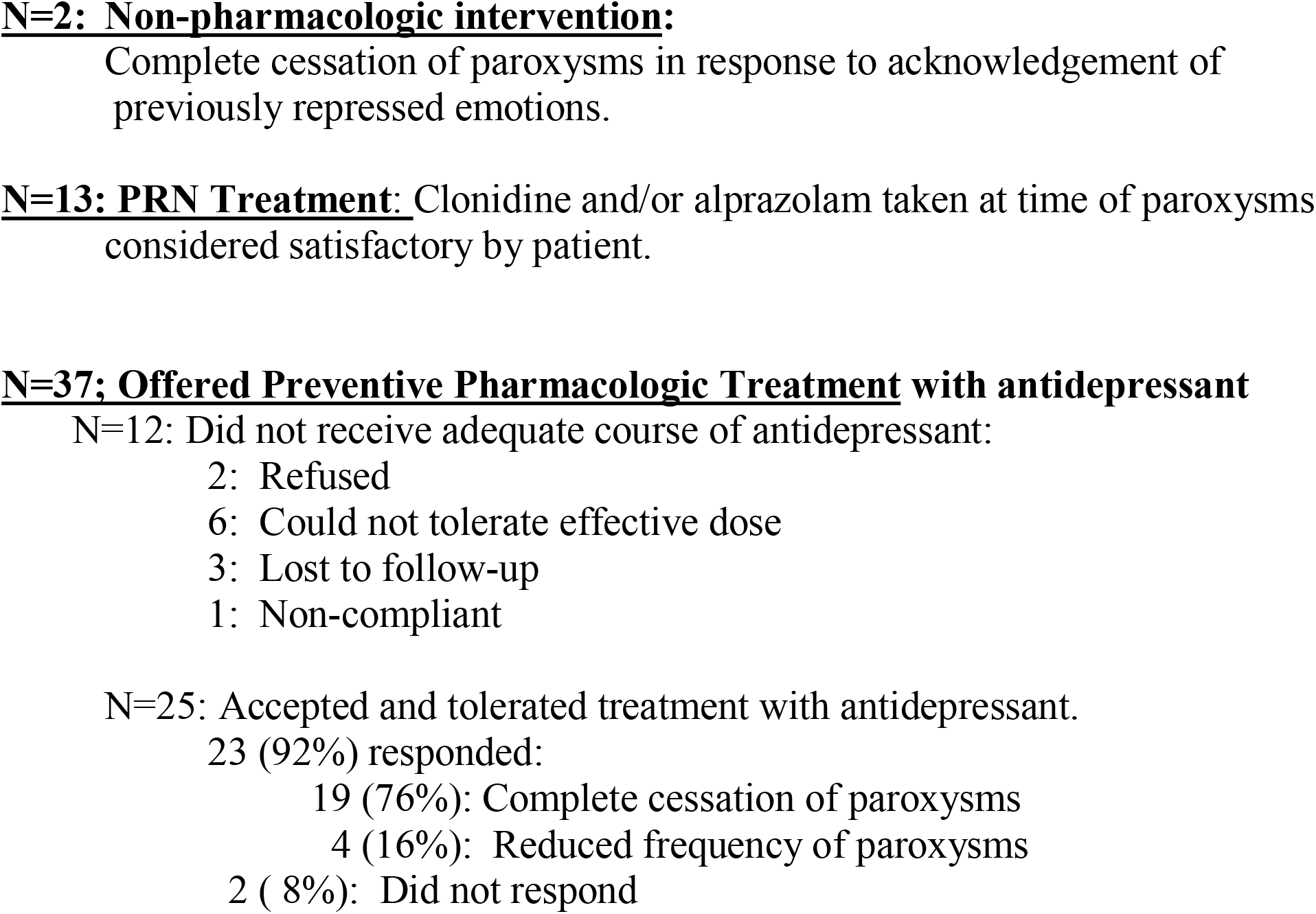
Summary: Results of Treatment (n=52)

Treatment with an antidepressant was offered to 37 patients (71%). Two refused, and five were lost to follow-up. The remaining 30 patients were treated with the following agents: escitalopram (50%), citalopram (16%), bupropion (13%), desipramine (13%) and sertraline (10%). Four patients could not tolerate an effective dose, and one was non-compliant, leaving 25 evaluable responses.

Of the 25 evaluable patients, the response rate to an antidepressant was 92% (23/25), with 21 (84%) experiencing complete cessation of paroxysms and 2 (8%) experiencing a substantial reduction in frequency of paroxysms. Follow-up exceeded 6 months in 16 of the 25 patients, one year in 14 patients, and 2 years in 9.

The response rate was 58% (7/12) among patients who reported current anxiety or depression, and, strikingly, 86% (13/15) among those who did not report current anxiety or depression. Information regarding self-reported anxiety or depression was not available in the charts of 3 patients.

Psychosocial history revealed a history of abuse, trauma, untimely bereavement, or severe family dysfunction in 17 of the 25 patients. Fifteen of the 17 responded to the antidepressant.

Finally, and of great interest, a prompt and persisting cessation of paroxysms without medication was observed in two patients who had been suffering paroxysms for at least 5 years. They did not report a history of trauma, but in both cases cessation of paroxysms followed the very emotional acknowledgement of emotions related to a past history of severe prolonged stress. One had divorced her husband in her 20s, followed by years of overwhelming responsibilities simultaneously raising two toddlers, holding two jobs and attending college. The other was an immigrant who had uncomplainingly worked 17-hour days, 7 days a week for the first two decades he was in this country.

## Discussion

### Management of paroxysmal hypertension

The high response rate to an antidepressant observed in this report is highly consistent with that observed in two previous reports.^3,4^ Even in the absence of a placebo-treated control group, the consistent very high response rate, now reported in three studies, conveys clearly that treatment with an antidepressant offers a pioneering, highly effective advance in the management of paroxysmal hypertension. Given the absence of evidence that any other treatment can prevent recurrence of paroxysms, and the disruptive effect of recurrent paroxysms, and the known relative safety of antidepressant agents, the results strongly support an empiric trial of an antidepressant in managing patients with severe or frequent paroxysms.

In this case series, antidepressant treatment to prevent paroxysms was offered to 71% of patients (37/52). Patients with relatively mild or infrequent paroxysms might not need the possibly lifelong treatment with an antidepressant.

### Acute management of paroxysms

In both this and the previous case series, clonidine or a benzodiazepine or a combination of the two was observed to lower blood pressure and reduce symptoms in nearly all patients, usually within 30 to 60 minutes.^3^ It was considered to be satisfactory and sufficient treatment by patients with relatively mild or infrequent paroxysms. The benzodiazepine alprazolam (0.25-0.5 mg) was often prescribed because of its rapid onset of effect. Importantly, patients are able to self-medicate at the onset of paroxysms, often obviating the need to repeatedly seek emergency room care.

The rapid increase in blood pressure strongly suggests the involvement of the sympathetic nervous system (SNS), as also suggested by the observed concomitant increase in plasma catecholamines .^6,7^ Consistent with this, intravenous administration of an effective dose of the alpha/beta-blocker labetalol is usually effective within 5-10 minutes. The onset of action of oral labetalol is usually within 60-90 minutes; however, the considerable inter-individual variance in the effective dose is problematic.

The role of oral agents that target the renin-angiotensin system (e.g. angiotensin-converting enzyme inhibitors (ACE inhibitors) or angiotensin receptor blockers (ARBs) or sodium/volume mechanisms (e.g. diuretics) has not been assessed, but such agents would seem less likely to be effective than agents that target the SNS. Rapid-acting oral nifedipine is no longer recommended due to risks associated with its use. Intravenous nicardipine constitutes another possible though unassessed alternative.

It is important to recognize the risk of hypotension following acute treatment of paroxysms. The risk is attributable to multiple factors including the antihypertensive effect of the acutely administered antihypertensive agent, interaction with previously taken antihypertensive medications, and volume loss resulting from diaphoresis and/or pressure natriuresis during paroxysms. In this regard, though it has never been assessed, it is possible that acute management with a benzodiazepine, if effective, might be safer than antihypertensive agents, particularly in patients who are normotensive between paroxysms.

### Chronic preventive management

#### Clinical indications for Prescribing an Antidepressant

If paroxysms are infrequent and not severe, medication taken as needed when paroxysms occur is often sufficient. However, if blood pressure elevation is severe, or if paroxysms are very symptomatic, frequent, terrifying to the patient, significantly interfering with functioning, or are occurring in a patient at increased risk of complications due to co-morbidities, treatment with an antidepressant to prevent paroxysms merits strong consideration.

#### Is there a Role for Preventive Treatment with Antihypertensive Agents?

In this case series, treatment with antihypertensive agents was not observed to prevent paroxysms. Chronic treatment with agents that target the SNS, such as alpha/beta-blockade, could be postulated to be more effective than treatment with agents that target volume or the RAS in reducing the peak blood pressure during paroxysms, but this possibility has not been studied. An additional concern in prescribing an antihypertensive regimen for chronic prevention in patients who are normotensive between paroxysms is the risk of hypotension following acute administration of additional antihypertensive agents during a paroxysm.

### Differential Diagnosis

The conditions that should be most prominently considered in the differential diagnosis of paroxysmal hypertension are *pheochromocytoma, labile hypertension*, and *panic disorder*. Other conditions that can also be associated with labile elevation of blood pressure include, but are not limited to, hypertensive encephalopathy, renovascular hypertension, clonidine withdrawal, MAO inhibitor crisis, migraine, cluster headache, carcinoid, mastocytosis, intracranial lesions, and baroreflex failure.

Although a *pheochromocytoma* is found in only 1-2% of patients with paroxysmal hypertension, a screening test, such as plasma metanephrines, should be performed given the serious consequences of missing the diagnosis. ^2^,^8^ Mild elevation of plasma metanephrines is not uncommon and is not indicative of a pheochromocytoma; a 30% false positive rate has been reported, and may be associated with lack of adherence to fasting state, supine position, and rest before sampling.^9^

To date there are no diagnostic criteria for the diagnosis of *labile hypertension*. Diagnosis is by clinical impression. Labile hypertension can be asymptomatic, or can be associated with symptoms such as headache and/or palpitations.^10^

Patients who meet the three proposed clinical criteria listed above are highly likely to have paroxysmal rather than labile hypertension. Importantly, nearly all patients with paroxysmal hypertension describe paroxysms as occurring “out of the blue,” and insist that the paroxysms are not related to anxiety or stress. In contrast, most patient with labile hypertension acknowledge feeling anxious or stressed at the time of blood pressure elevation, and most attribute the blood pressure elevation to the emotional distress. Distinguishing between the two is important because of the effectiveness, and often the necessity, of treatment with an antidepressant in patients with paroxysmal hypertension.

*Panic disorder* resembles paroxysmal hypertension in that both manifest with sudden, unprovoked attacks and similar physical symptoms. They differ in that panic attacks are usually dominated by panic and are accompanied by a lesser degree of blood pressure elevation, whereas paroxysmal hypertension is usually dominated by marked blood pressure elevation.^11^ Also, panic attacks are heralded by the experience of panic; in patients with paroxysmal hypertension, the fear and anxiety occur instead as a reaction to the frightening physical symptoms.

## Exploring a Possible Psychosomatic Etiology

The effectiveness of treatment with an antidepressant strongly suggests a psychosomatic etiology. Yet a psychosomatic etiology is rarely suspected because patients uniformly insist that their paroxysms are *not* related to current stress or emotional distress. Further, most patients in this report did not report current anxiety or depression.

However, in this case series as well as in the earlier series, a past history of significant trauma, abuse, untimely death of a parent or spouse, or other overwhelming stress was noted in most patients.^3^ Such events typically are not uncovered when psychosocial inquiry focuses only on current stress or emotional distress.

The absence of reported current emotional distress and the insistence by many patients that past trauma had had no lingering emotional impact, along with the response to antidepressants, suggest the possible role of emotions that were of necessity repressed, as a means of coping, at the time of severe stress or trauma. Similarly, the claim by some patients of “never” experiencing symptoms of anxiety or depression despite a stressful life history suggests a “repressive coping style.” ^12^ Such patients would otherwise be unlikely to be offered treatment with an antidepressant.

This psychosomatic understanding is also strongly supported by the prompt resolution of the disorder, without medication, observed in two patients after they gained awareness of previously repressed emotions related to decades-old overwhelming stress. This observation strongly suggests that in resilient survivors of overwhelming stress or trauma, a core of repressed emotions can and does persist.

The potential benefit of psychotherapy in this disorder is not known. The absence of emotional distress and the powerful barrier posed by repression understandably constitute a significant impediment to effective psychotherapy. Fortunately, treatment with an antidepressant provides a very effective alternative.

### Limitations

To date a controlled trial comparing an antidepressant versus a placebo has not been performed. However, the very high response rate in this and in previous reports far exceeds any expected placebo response.^3,4^ It is possible that some of the patients who were lost to follow-up might have been non-responders, but even if that were the case, the response rate nevertheless remains very high.

Another limitation is that this report concerns patients whose accrual occurred sporadically, in a clinical rather than a study setting. The history of trauma or overwhelming stress was ascertained by careful clinical history without the use of a formal trauma questionnaire. However, this limitation should not negate the findings. Trauma questionnaires such as the widely employed Adverse Childhood Experiences Survey (ACES) possess important weaknesses compared to careful clinical history-taking.^13^ The “yes” or “no” format for responses ignores the details that determine the severity or lack of severity of any given “trauma.” Also, the derived trauma score is based on the number of positive answers; it ignores the severity of any single event. Finally, the ACES questionnaire omits major events such as the death or loss of a parent during childhood, and does not take into consideration trauma that occurred in adulthood. For these reasons, crucial information obtained by clinical history, particularly as it pertains to potentially traumatic events, must not be ignored.

## Conclusions

This is the third report documenting the pioneering finding that treatment with an antidepressant is uniquely effective in preventing recurrent paroxysms in most patients with paroxysmal hypertension.^3,4^ The high response rate suggests a psychosomatic origin despite the absence, in most patients, of perceived emotional distress. A psychosomatic origin is also strongly supported by observation of complete resolution of the disorder without any medication in two patients after acknowledgment of long-repressed emotions.

The three main treatment options are: (1) treatment as needed when paroxysms occur with a benzodiazepine and/or clonidine, (2) ongoing treatment with an antidepressant to prevent paroxysms, and (3) in a minority of patients, psychological healing that can occur with acknowledgement of long-repressed painful emotion. There is no reported evidence that antihypertensive agents can prevent paroxysms.

To date no alternative understanding or treatment options exist. More widespread attention to the effectiveness of antidepressants is needed, particularly given the severe symptoms and chronicity, the disruption to patients’ lives, and the avoidable emergency room visits and hospitalizations associated with this disorder.

## Data Availability

Data collected from patient medical records that was included in this paper are available for review.

